# Substantial reduction in the clinical and economic burden of disease following variant-adapted mRNA COVID-19 vaccines in immunocompromised patients in France

**DOI:** 10.1101/2024.03.13.24304170

**Authors:** Amy Lee, Benjamin Davido, Ekkehard Beck, Clarisse Demont, Keya Joshi, Michele Kohli, Michael Maschio, Mathieu Uhart, Nadia El Mouaddin

## Abstract

An economic evaluation was conducted to predict the economic and clinical burden of vaccinating immunocompromised (IC) individuals aged ≥30 years with mRNA-1273 variant-adapted COVID-19 vaccines in Fall 2023 and Spring 2024 versus BNT162b2 variant-adapted vaccines in France. The number of symptomatic COVID-19 infections, hospitalizations, deaths, and long COVID cases, costs and quality-adjusted life years (QALYs) was estimated using a static decision-analytic model. Predicted vaccine effectiveness (VE) were based on real-world data from prior versions, suggesting higher protection against infection and hospitalization with mRNA-1273 vaccines. VE estimates were combined with COVID-19 incidence and probability of COVID-19 severe outcomes. Uncertainty surrounding VE, vaccine coverage, infection incidence, hospitalization and mortality rates, costs and QALYs were tested in sensitivity analyses. The mRNA-1273 variant-adapted vaccine is predicted to prevent an additional 3,882 infections, 357 hospitalizations, 81 deaths, and 326 long COVID cases when compared to BNT162b2 variant-adapted vaccines in 230,000 IC individuals. This translates to €10.1 million cost-savings from a societal perspective and 645 QALYs saved. Results were consistent across all analyses and most sensitive to variations surrounding VE and coverage. These findings highlight the importance of increasing vaccine coverage, and ability to induce higher levels of protection with mRNA-1273 formulations in this vulnerable population.

## INTRODUCTION

In December 2020, France launched its first COVID-19 vaccination campaign against the SARS-CoV-2 virus [1], with priority vaccination of the most vulnerable populations, including elderly and immunocompromised (IC) individuals [2, 3]. As of the beginning of October 2023, over 160.4 million doses have been administered in France, with 78.7% of the population completing the primary course of vaccinations with one of the recommended vaccines in France [4].

Severely IC individuals (solid organ transplant recipients, recent bone marrow transplant recipients, dialysis patients, patients with autoimmune disease receiving immunosuppressive treatments, patients receiving treatment with anti-CD20 [including certain types of lymphoma] or antimetabolites, and chronic lymphocytic leukemia patients) are at increased risk of severe outcomes following COVID-19 infection [5]. In France, 43% of those hospitalized in critical care due to the SARS-CoV-2 virus (Omicron variant) between December 2021 and May 2022 were IC individuals [6].

The protective response to vaccines mounted by IC individuals has been found to be lower compared to the general population. For example, in a study of Kaiser Permanente Southern California patients receiving the mRNA-1273 (Moderna Spikevax) BA.4/BA.5 variant-adapted bivalent vaccine, Tseng et al., (2023) found the adjusted vaccine effectiveness (VE) against hospitalization for the general population to be 82.8% compared to 71.8% in those that were IC [7].

Additionally, despite IC patients not being enrolled in the pivotal Phase 3 COVID-19 clinical trials, a recent 2024 mRNA-1273 update by the European Medicines Agency (EMA) now includes clinical trial data on safety and immunogenicity for solid organ transplant patients [8]. These newly available data may also help to address potential concerns of the IC population with regards to safety and efficacy [9, 10].

Although both mRNA-1273 and BNT162b2 (Pfizer-BioNTech Comirnaty) COVID-19 vaccines and their subsequent variant-adapted vaccines are based on mRNA technology, the formulations of mRNA-1273 version, including their dosages and nanoparticle mRNA delivery systems, differ [11–14]. Studies have found VE differences between previous versions of the mRNA-1273 vaccine and the corresponding BNT162b2 vaccine in both the general [15–19] and IC populations [20], with mRNA-1273 formulations resulting in greater levels of protection. Wang et al., (2023) conducted a systematic literature review and meta-analysis based on 17 studies in the IC population [20]. The mRNA-1273 vaccine was found to be associated with a significantly lower risk of COVID-19 infection (relative risk [RR] for infection = 0.85 [95% CI 0.75-0.97]), hospitalizations (RR = 0.88 [95% CI 0.79-0.97]), and deaths (RR = 0.63 [95% CI 0.44-0.90]), compared to the BNT162b2 vaccine.

In May 2023, the World Health Organization (WHO) declared that the pandemic is no longer a public health emergency of international concern, but stated that the disease still requires ongoing management [21]. The French Ministry of Health and National Authority for Health namely the Haute Autorité de Santé (HAS) recommended that the Fall 2023 and the Spring 2024 vaccination campaigns target those at higher risk of severe outcomes from the disease, including the IC population [22–24], which are estimated to comprise approximately 230,000 individuals in France [5].

The French Ministry of Health and HAS further recommended use of mRNA variant-updated vaccines to the dominant circulating strain (XBB.1.5) [22, 23]. Although mRNA-1273 vaccine formulations are deemed to produce higher antibodies responses than their BNT162b2 counterparts [25–27], which is important especially for this targeted high-risk population, and while both mRNA-1273 and BNT162b2 monovalent variant-adapted (XBB.1.5) formulations are approved for use in France, only the BNT162b2 XBB.1.5 variant-adapted version was available in France for the Fall 2023 vaccination campaign.

Having the choice between two vaccines will fill a void, providing the vulnerable IC population with the option of a vaccine that offers higher levels of protection. Additionally, as the XBB.1.5 variant-adapted formulation of mRNA-1273 is packaged as single-use pre-filled syringe, having the option of individual doses instead of multi-dose vials could reduce vaccine wastage due to unfinished, opened vials and could therefore increase the availability of vaccines for others world-wide [28, 29].

Thus, this study aims to estimate the maximum public health and economic impact by vaccinating with mRNA-1273 variant-adapted vaccines in Fall 2023 and Spring 2024 compared to vaccinating with BNT162b2 variant-adapted vaccines during the same time period. The differential impact on clinical outcomes and health care costs in IC individuals over a one-year time horizon between October 2023 and September 2024 is examined to accurately reflect the burden of disease due to COVID-19 in this vulnerable population.

## METHODS

### Overview

As prior versions of the mRNA-1273 COVID-19 vaccines in France were targeted to adults aged 30 years and older, in accordance with HAS guidelines [23], the target population of this analysis is also IC individuals aged 30 years and older in France.

A static decision analytic model, previously used on the Canadian IC population [30], was adapted to France using country-specific inputs. Full details of the model are described by Lee et al., (2023) [30], and a brief description is provided here. In the Markov model, the monthly COVID-19 vaccination status of the target population of IC individuals aged 30 years and older in France are tracked with the use of tunnel states. The model was developed in Microsoft Excel, and estimates the clinical (infections, hospitalizations, deaths, quality-adjusted life-years [QALYs]) and costs associated with COVID-19 infection treatment over a one-year time horizon (October 2023 – September 2024), taking into account France-specific estimates of COVID-19 incidence, the effectiveness of prior versions of COVID-19 mRNA vaccinations and the probability of severe outcomes from COVID-19 infections. Economic costs were estimated from both healthcare and societal perspectives.

In the model, proportions of the IC population aged 30 years and above are assigned to one of five historical vaccination strata, based on the highest level of vaccination received prior to the start of the time horizon (October 2023): did not complete primary series; primary series (2 doses); first booster; second booster, third booster. Data on COVID-19 vaccine coverage in the general population were obtained from the European Centre for Disease Control (ECDC), which included vaccine doses delivered for each vaccination strata by age group [4], and used for the IC population.

It is difficult to estimate the vaccine coverage rate in the IC population, as hospital administered COVID-19 vaccines are not reported in the French National Health Data System (SNDS) and ECDC data on COVID-19 coverage are only reported for general population. For the base case analysis, an ideal scenario with 100% of the IC population aged 30 years and above receiving a mRNA-1273 variant-adapted Fall 2023 vaccine in October 2023 and a Spring 2024 vaccine in April 2024 was assumed. This was compared to outcomes where 100% of the IC population received BNT162b2 variant-adapted Fall 2023 and Spring 2024 vaccines. Several scenario analyses were also conducted, with lower than 100% vaccine coverage for both mRNA-1273 and BNT162b2 versions of the vaccines. In the first scenario analysis (“alternate”), it was considered that 50% of the IC population received the Spring 2024 vaccine. A second scenario (“flu-like”) was included where those aged 65 years and over assumed a coverage rate equal to that observed during the 2021-22 influenza vaccination campaign in France (56%) [31], and those aged 30-64 years old assumed the same coverage as that reported for influenza vaccination in IC gastroenterology patients (20%) [32]. Finally, a third coverage scenario analysis (“pessimistic”) was included where those aged 65 years and over assumed the coverage as that observed for influenza vaccination in IC gastroenterology patients (20%), and those aged 30-64 years assumed the same coverage, by age, as that observed in France with the second COVID-19 booster (3.2% for ages 30-49 years, 11.7% for ages 50-64 years) [33].

To determine the incidence and level of vaccine-induced protection of the population entering the decision analytic model, a separate dynamic Susceptible-Exposed-Infected-Recovered (SEIR) model was used to calculate the changes in incidence and protection levels over time. This model, which has been described in detail in a published US cost-effectiveness analysis [34], was adapted with French-specific inputs, including population size, prior vaccination coverage, and historical incidence. The model predicted residual VE of the French general population, from prior vaccinations, as well as the infection incidence from October 2023 to September 2024. Although for the base case, 100% of individuals are vaccinated in Month 1 and therefore assumes the VE of the Fall 2023 vaccine at the start of the analytic time horizon, residual VE is still important for the portions of the population that do not immediately receive the Fall 2023 vaccine in the coverage scenario analyses. See the Technical Appendix for further details.

At the start of the analytic time horizon, the cohort enters the model in the Well health state (Figure 1), where they are at risk of COVID-19 symptomatic infection. This risk depends on the monthly incidence of COVID-19 symptomatic infection, and is modified by their prior vaccination status and whether they received a Fall 2023 and Spring 2024 vaccine. Individuals that develop a COVID-19 infection then move through an infection consequence decision tree (Figure 2, described in detail further below), which calculates clinical outcomes and costs associated with the infection. Individuals remain in the Well health state (shown as Recovered in the consequence decision tree) unless they experience death in hospital due to the COVID-19 infection (shown as In-Hospital Mortality in the decision tree, and Death in the Markov model). Death in the model is only due to COVID-19 due to the short time horizon. All-cause mortality is not included.

**Figure 1.**
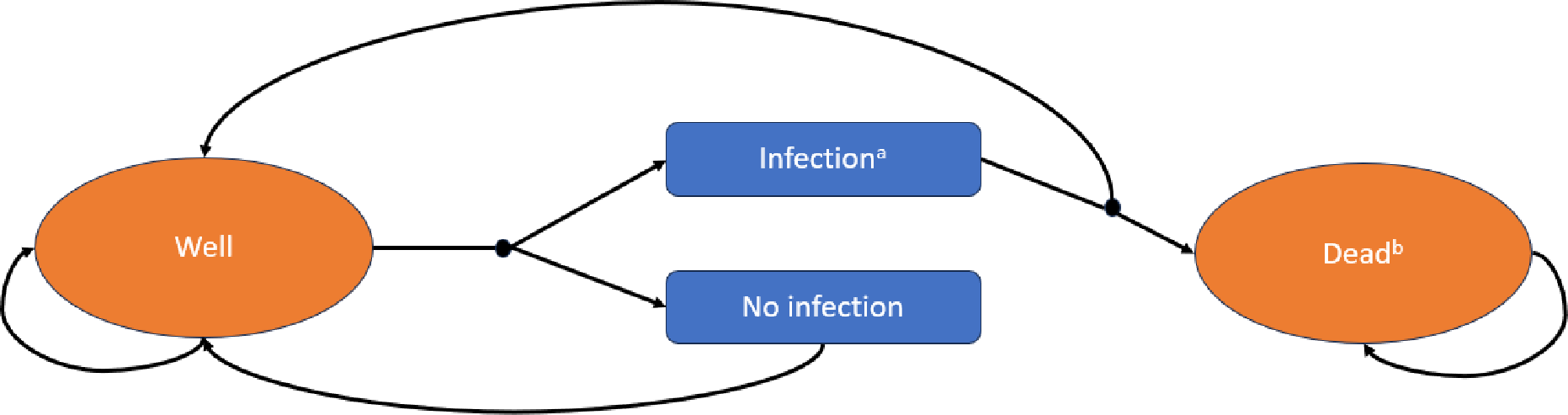
Model structure. ^a^ see infection consequence decision tree structure (Figure 2) ^b^ death due to COVID-19

**Figure 2.**
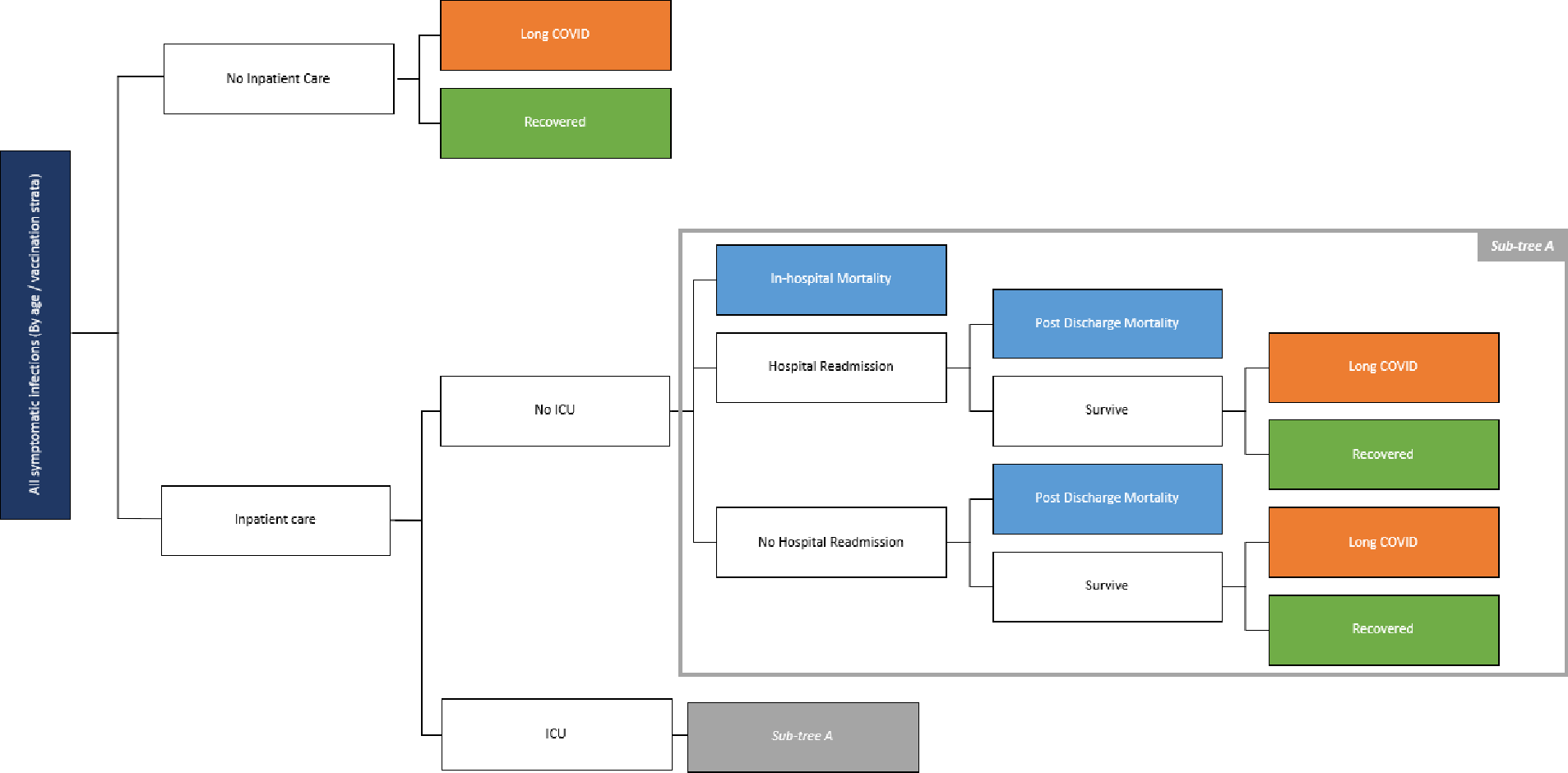
Infection consequence decision tree structure.

### Population size

It is estimated that approximately 230,000 individuals in France are IC [35]. As French data on the IC population distribution by age were not available, Canadian data were used. Ramage-Morin and Polsky conducted a survey during the initial months of the COVID-19 pandemic to determine the proportion of individuals, by age group, that were immunocompromised [36]. These proportions were reweighted by the French general population distribution, and then multiplied by 230,000 to estimate the IC population size for each model age group. French general population estimates for 2020 (which was used to generate monthly incidence in the dynamic SEIR model, as the SEIR model simulations begin in the year 2020) were obtained from the United Nations Department of Economic and Social Affairs[37]. The 230,000-population size was applied to individuals aged 30 years and over, although the estimate may have contained children and young adults as well. Resulting data by age are displayed below in Table 1.

**Table 1.**
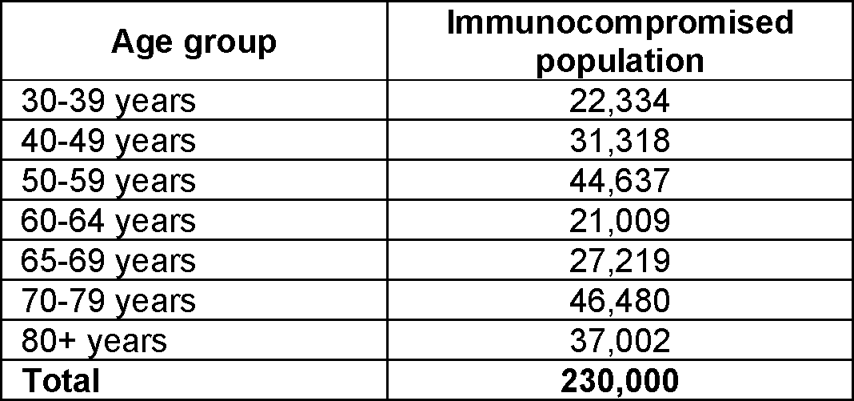
French population aged 30 years and over, by age.

### Incidence of symptomatic infection

The projected monthly COVID-19 symptomatic infection incidence rates without vaccination for October 1, 2023 to September 30, 2024 (the next fall season), during the analytic time period is age-specific and as previously mentioned, was predicted using the dynamic SEIR model. Methodology is described in detail in the US cost-effectiveness analysis publication [34], and parameters adapted to France are listed in the Technical Appendix. The resulting projected monthly incidence of symptomatic COVID-19 symptomatic infection among the unvaccinated in France in the model is displayed in Table 2.

**Table 2.** Projected incidence of symptomatic COVID-19 infection amongst the unvaccinated in France (%)

| Age Group | Month* |  |  |  |  |  |  |  |  |  |  |  |
| --- | --- | --- | --- | --- | --- | --- | --- | --- | --- | --- | --- | --- |
|  | 1 | 2 | 3 | 4 | 5 | 6 | 7 | 8 | 9 | 10 | 11 | 12 |
| 30-39 years | 1.77 | 2.12 | 2.69 | 2.76 | 2.22 | 1.95 | 1.66 | 1.69 | 1.75 | 2.00 | 2.20 | 2.27 |
| 40-49 years | 1.76 | 2.10 | 2.67 | 2.76 | 2.22 | 1.95 | 1.66 | 1.68 | 1.73 | 1.98 | 2.19 | 2.26 |
| 50-59 years | 1.66 | 1.97 | 2.55 | 2.72 | 2.23 | 1.94 | 1.60 | 1.58 | 1.60 | 1.82 | 2.03 | 2.13 |
| 60-64 years | 1.51 | 1.74 | 2.29 | 2.56 | 2.18 | 1.88 | 1.49 | 1.40 | 1.36 | 1.52 | 1.69 | 1.80 |
| 65-69 years | 1.51 | 1.74 | 2.29 | 2.55 | 2.18 | 1.88 | 1.49 | 1.40 | 1.36 | 1.52 | 1.69 | 1.80 |
| 70-79 years | 1.46 | 1.67 | 2.19 | 2.48 | 2.13 | 1.84 | 1.45 | 1.34 | 1.29 | 1.42 | 1.58 | 1.68 |
| ≥80 years | 1.41 | 1.61 | 2.13 | 2.41 | 2.07 | 1.79 | 1.41 | 1.30 | 1.25 | 1.39 | 1.53 | 1.63 |

\*Month 1 = October 2023.

A lower symptomatic infection incidence scenario, where the waning of naturally acquired immunity was assumed to be slower than the base case, was tested in a scenario analysis, to explore the impact of incidence on outcomes. Details on the calculation of the monthly incidence are provided in the Technical Appendix. Lower and higher symptomatic infection incidence scenario, where the base case value is varied by ±25%, were also explored.

### Vaccine effectiveness of the mRNA-1273 and BNT162b2 variant-adapted Fall 2023 and Spring 2024 vaccines

Real-world effectiveness specific to mRNA-1273 variant-adapted and BNT162b2 variant-adapted Fall 2023 and Spring 2024 vaccines are not yet available. In this analysis, both Fall 2023 and Spring 2024 vaccines are assumed to have the same level of protection against circulating strains as observed with previous versions. Using recent data, the mRNA-1273 variant-adapted Fall 2023 and Spring 2024 vaccines are assumed to have the same VE against hospitalization in the IC population as the mRNA-1273 bivalent vaccine (mRNA-1273.222) against BA.4/BA.5 (71.8%) in the IC population [38]. As the absolute VE against infection for the mRNA-1273 bivalent and XBB1.5 vaccine is not available, the mRNA-1273 variant-adapted Fall 2023 and Spring 2024 VE against infection was estimated from a meta-analysis [39] of real-world data on the original monovalent vaccine (mRNA-1273) against BA.1/BA.2 (57.1%). This value was based on the general population as IC data or adjustments are unavailable. This proposed value is plausible, as early estimates on XBB.1.5 variant-adapted vaccines (both mRNA-1273 and BNT162b2 combined) from the United States found the relative VE against infection of those that received the updated vaccines to be 54% compared to those that did not receive the updated vaccines [40]. Assuming a population fully waned against infection protection, this approximates the absolute VE, which is similar to our assumption of 57.1%. Additionally, an early Danish study on both mRNA-1273 and BNT162b2 XBB.1.5 variant adapted vaccines found a 71.6% reduction in hospitalization risk in those that received the updated vaccine, compared to those that only received the bivalent formulations a year prior [41].

The VE in the IC population for the BNT162b2 variant-adapted Fall 2023 and Spring 2024 vaccines were calculated using the assumed VE values for the mRNA-1273 variant-adapted Fall 2023 and Spring 2024 vaccines and the relative risk (RR) of hospitalization (RR=0.88 95% CI 0.79-0.97) and infection (RR=0.85, 95% CI 0.75-0.97) between mRNA-1273 and BNT162b2 vaccines from a pairwise meta-analysis in IC individuals [20]. Monthly waning for both vaccines are assumed to be the equivalent and based on a meta-analysis on the duration of vaccine protection [42]. Kopel et al., (2024) conducted a retrospective cohort study and estimated the VE of mRNA-1273 relative to BNT162b2 BA.4/BA.5 bivalent vaccines in the IC population against outpatient (relative VE = 15.0% [95% CI 7.2%-22.0%]; p<0.001) and hospitalizations (relative VE = 4.8% [95% CI 1.8%-7.7%]; p=0.020) [43]. These values were applied to the base case mRNA-1273 variant-adapted Fall 2023 and Spring 2024 vaccine VE values, with the outpatient relative VE as a proxy for infection, to estimate sensitivity analysis values for BNT162b2 variant-adapted Fall 2023 and Spring 2024 values. Assumed VEs and monthly waning rates for both mRNA-1273 and BNT162b2 variant-adapted Fall 2023 and Spring 2024 vaccines, as well as values used in sensitivity analyses based on VE 95% CIs are displayed in Table 3.

**Table 3.**
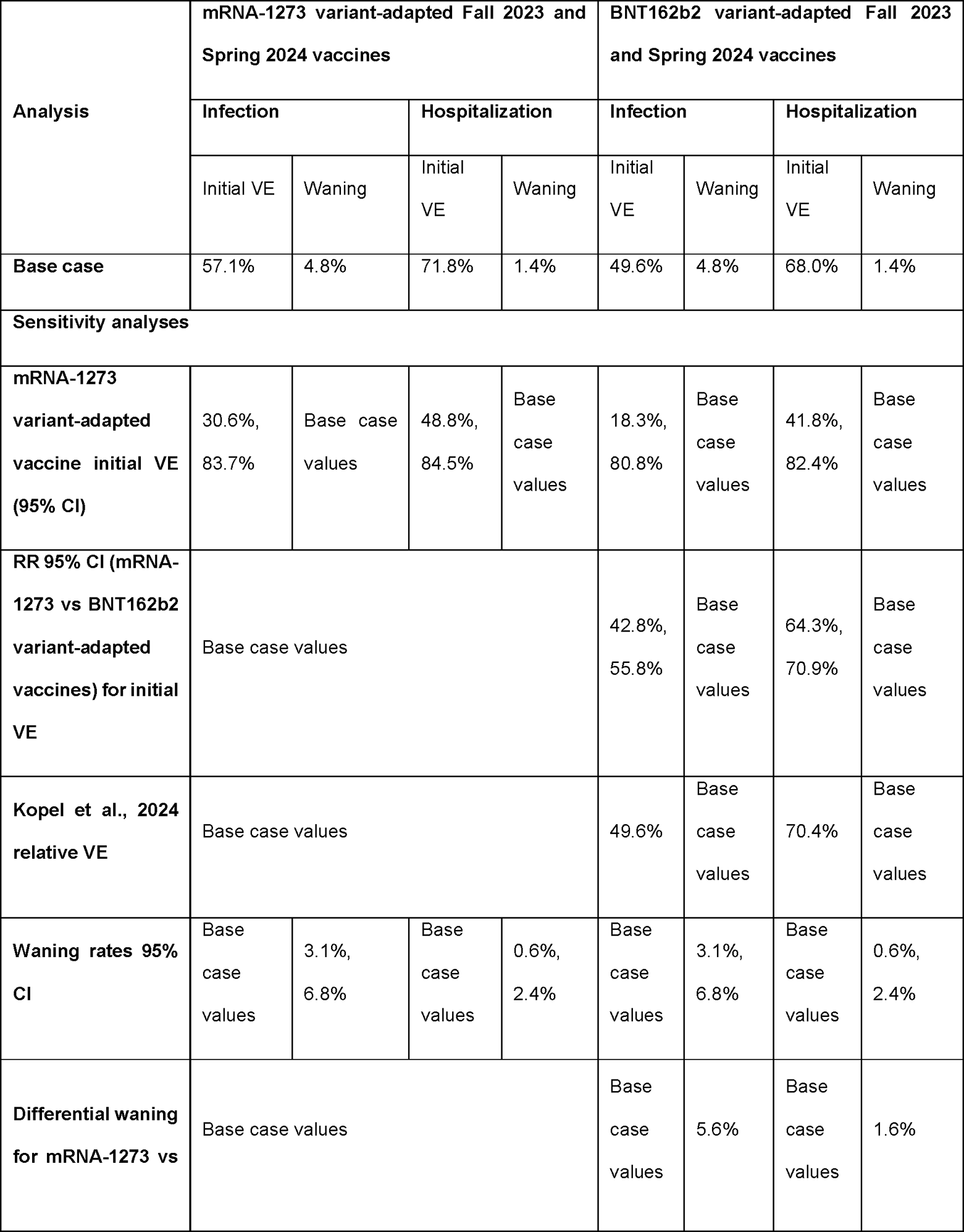
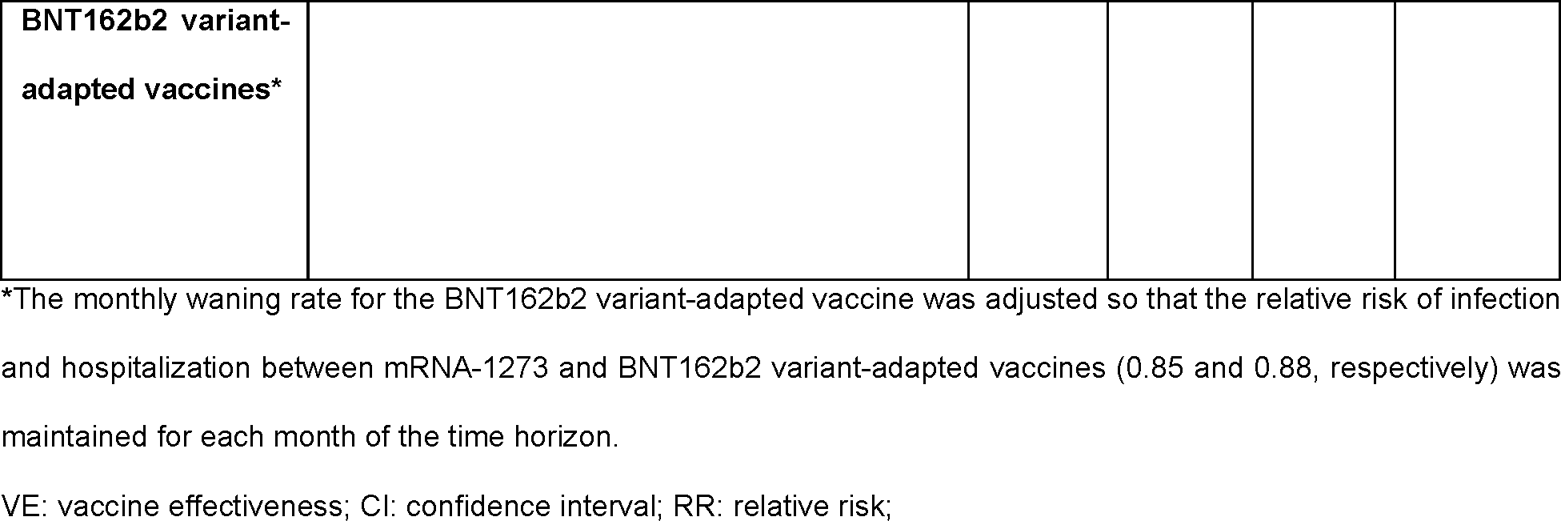
Base case and sensitivity analyses vaccine effectiveness inputs.

| Analysis | mRNA-1273 variant-adapted Fall 2023 and Spring 2024 vaccines |  |  |  | BNT162b2 variant-adapted Fall 2023 and Spring 2024 vaccines |  |  |  |
| --- | --- | --- | --- | --- | --- | --- | --- | --- |
|  | Infection |  | Hospitalization |  | Infection |  | Hospitalization |  |
|  | Initial VE | Waning | Initial VE | Waning | Initial VE | Waning | Initial VE | Waning |
| **Base case** | 57.1% | 4.8% | 71.8% | 1.4% | 49.6% | 4.8% | 68.0% | 1.4% |
| **Sensitivity analyses** |  |  |  |  |  |  |  |  |
| **mRNA-1273 variant-adapted vaccine initial VE (95% CI)** | 30.6%,<br>83.7% | Base case values | 48.8%,<br>84.5% | Base case values | 18.3%,<br>80.8% | Base case values | 41.8%,<br>82.4% | Base case values |
| **RR 95% CI (mRNA-1273 vs BNT162b2 variant-adapted vaccines) for initial VE** | Base case values |  |  |  | 42.8%,<br>55.8% | Base case values | 64.3%,<br>70.9% | Base case values |
| **Kopel et al., 2024 relative VE** | Base case values |  |  |  | 49.6% | Base case values | 70.4% | Base case values |
| **Waning rates 95% CI** | Base case values | 3.1%,<br>6.8% | Base case values | 0.6%,<br>2.4% | Base case values | 3.1%,<br>6.8% | Base case values | 0.6%,<br>2.4% |
| **Differential waning for mRNA-1273 vs** | Base case values |  |  |  | Base case values | 5.6% | Base case values | 1.6% |

|  |
| --- |
| **BNT162b2 variant-adapted vaccines\*** |

\*The monthly waning rate for the BNT162b2 variant-adapted vaccine was adjusted so that the relative risk of infection and hospitalization between mRNA-1273 and BNT162b2 variant-adapted vaccines (0.85 and 0.88, respectively) was maintained for each month of the time horizon.
VE: vaccine effectiveness; CI: confidence interval; RR: relative risk;

### Infection consequences decision tree

The infection consequence decision tree estimates the occurrences of the following health outcomes over the one-year analytic time horizon: cases of symptomatic infections, hospitalizations, COVID-19 related deaths, infection-related myocarditis (not shown in Figure 2), and long COVID. The decision tree also sums QALY losses that occur due to the listed health outcomes. Included cost categories are acute infection costs (outpatient visits and hospitalizations), infection-related myocarditis, long COVID, and for the societal perspective, productivity losses. As adverse events are expected to be similar between the mRNA-1273 and BNT162b2 variant-adapted vaccines, they were not included. The expected numbers of life-years lost due to early deaths from COVID-19 are calculated using expected survival by age. These data were obtained from the French National Institute of Statistics and Economic Studies [44]. The difference between the age at death due to COVID-19 and the age-specific expected survival are calculated by the model. Age-specific utility values for individuals without infection are attached to each year lost due to early death from COVID-19.

Transition probabilities used in the consequences decision tree (Figure 2), are displayed in Table 4. Data inputs were sourced from targeted literature reviews and were estimated based on published sources, fee schedules, and databases. Where available, French-specific sources and data were used.

**Table 4.** Base case infection consequences decision tree probabilities.

| Age group (years) | Hospitalization rates (unvaccinated) given infection, by age* | Hospitalization locations of care, by age |  | In-hospital mortality rates, by age* | Excess risk of infection-induced myocarditis, by age [45] |
| --- | --- | --- | --- | --- | --- |
|  |  | General ward | ICU |  |  |
| 30-39 | 4.62% | 92.80% | 7.20% | 1.56% | 0.07% |
| 40-49 | 4.52% | 76.24% | 23.76% | 6.96% | 0.09% |
| 50-59 | 4.52% | 76.24% | 23.76% | 6.96% | 0.14% |
| 60-64 | 22.03% | 77.32% | 22.68% | 18.75% | 0.15% |
| 65-69 | 22.03% | 77.32% | 22.68% | 18.75% | 0.16% |
| 70-79 | 22.03% | 77.32% | 22.68% | 18.75% | 0.19% |
| 80+ | 51.63% | 94.47% | 5.53% | 26.62% | 0.21% |

\* Adjusted for immunocompromised status[49, 51]
ICU: Intensive care unit

Not all non-hospitalized patients were assumed to seek care. The cost per outpatient care was only applied to the proportion assumed to seek care, which was estimated from influenza data, adjusted by population by age in France (25.1%) [37, 46, 47]. However, the QALY loss for non-hospitalized patients was applied as a toll to all non-hospitalized patients whether care was sought or not.

A proportion of infected individuals require inpatient care (hospitalization). Risk of hospitalization in those without COVID-19 vaccine protection is expressed as the risk of hospitalization given infection in the unvaccinated population. Age-specific hospitalization rates in the unvaccinated population, given infection, was obtained from Direction de la Recherche, des Études, de l’Évaluation et des Statistiques (DREES) [48]. These values were based on the general population and were adjusted to reflect the IC population using a study by Bahremand et al., (2023) [49]. In this population-based observation study, individuals were categorized based on pre-existing health conditions. Data from groups considered to be IC were compared to the non-IC group, and a relative risk of hospitalization between the groups was calculated (RR=1.94). This value was applied to the hospitalization rates in the unvaccinated general population to approximate hospitalization rates in the IC (Table 4). The proportion of those hospitalized requiring an intensive care unit (ICU) stay (including those that required ventilation) was obtained from DREES [48]. It was assumed that if a hospitalized individual did not require an ICU stay, they were treated in the general wards. Individuals also were assigned a risk of hospital readmission (4.1%) [50] with costs the same as the initial admission. A hospitalization recovery cost was added to the unit cost of all hospitalizations. QALY losses due to hospitalizations were applied as a toll, with a different value applied to the proportion that were in the general ward vs. ICU, and with re-admission. Productivity loss inputs are provided in the Technical Appendix.

It was assumed that only hospitalized patients were at risk for COVID-19 related mortality. Age-specific mortality rates for those hospitalized for the French general population was obtained from DREES[48]. These values were adjusted to the IC population using data from a study by Turtle et al., (2023) [51] that compared the outcomes of COVID-19 hospitalized IC patients with immunocompetent patients (odds ratio (OR) for mortality in the IC group = 1.44 [95% CI 1.39, 1.5]). This increased odd of mortality is applied to the initial hospitalization. Individuals are also subject to post-discharge mortality (2.7%) [50] which is applied to both initial hospitalization and readmissions. As data were not available on the RR of post-discharge mortality in IC individuals compared to the general population, this value was not adjusted.

The QALY loss associated with early mortality due to COVID-19 was estimated by first calculating the expected number of life years lost [52]. Age-specific utility values for individuals without infection were attached to each year lost. All future QALYs lost were discounted at a rate of 2.5% to present value, as recommended in French Health Authorities guidelines [53].

Following the acute infection period, individuals are at risk of long COVID (also known as post-acute sequelae of SARS-CoV-2 infection (PASC)). Due to the lack of age-specific data, this risk (8% for non-hospitalized individuals) [54] was applied to all model age groups. For hospitalized individuals, the risk was assumed to be doubled (16%). Healthcare cost associated with long COVID was estimated through expert opinion, assuming a 6-month duration of symptoms [55–57]. Prevalence of long COVID, and its associated healthcare costs and QALY decrements are provided in Table 5.

**Table 5.** Base case infection consequences decision tree costs and QALY decrements.

| Model Parameter | Value | Data Source |
| --- | --- | --- |
| Infection-induced myocarditis cost | 1,802.41 € | Scansanté[58] / Tarifs GHS 2022 , Rapport cour des comptes[59] |
| QALY loss for infection-induced myocarditis | 0.0019 | Prosser et al., (2019)[60] |
| Non-hospitalized patients |  |  |
| Proportion seeking outpatient care | 25.1% | Réseau Sentinelle[47], CDC[46] |
| Cost per outpatient care | 277.73 € | Base des médicaments et Informations Tarifaires[55], Ameli,[56], Liste LPP, CCAM, Ministère de la santé |
| QALY loss | 0.0036 | Smith & Roberts (2002),[61] |
| Hospitalization cost (per stay) |  |  |
| General ward (no ICU) | 4,770.11 € | Gallien et al., (2020)[62, 63] |
| ICU | 17,092.71 € | Gallien et al., (2020)[62, 63] |
| Hospitalization recovery cost | 474.14 € | HAS. Avis économique Evusheld[64] |
| QALY loss |  |  |
| General ward (no ICU) | 0.0216 | Padula et al., (2021)[65] |
| ICU | 0.0466 | Padula et al., (2021)[65] |
| Hospital readmission | 0.0216 | Padula et al., (2021)[65] |
| Utility values (general population) |  |  |
| 30-39 years | 0.9290 | Szende et al., (2022)[66] |
| 40-49 years | 0.9174 |  |
| 50-59 years | 0.8881 |  |
| 60-64 years | 0.8530 |  |
| 65-69 years | 0.8100 |  |
| 70-79 years | 0.7609 |  |
| 80+ years | 0.7350 |  |
| Prevalence of long COVID<br>(both patients who were non-hospitalized and hospitalized) | 8% | Santé Publique France[54] |
| Long COVID costs | 917.71 € | Assurance Maladie, Official journal [55-57] |
| Long COVID QALY loss |  |  |
| Non-hospitalized | 0.0685 | Van Wilder et al., (2022)[67] |
| Hospitalized | 0.1390 | Van Wilder et al., (2022)[67] |

QALY, quality-adjusted life-year; ICU, intensive care unit; DREES, Direction de la recherche, des études, de l'évaluation et des statistiques

### Scenario analyses and deterministic sensitivity analyses

In addition to the scenario/sensitivity analyses already listed, analyses were conducted on hospitalization and mortality rates, costs, and QALY decrements. For the sensitivity analyses, lower and upper 95% CIs were used to define ranges; where unavailable, values were varied by ± 25%. For the proportion of infections that resulted in long COVID, sensitivity analyses were included where the proportion was set at 0% (i.e. no long COVID), as well as earlier estimates of 30% for non-hospitalized and 38% for hospitalized from Santé Publique France [68]. Value used for the included sensitivity analyses are presented in the Technical Appendix.

## RESULTS

Assuming vaccination of the entire IC population age 30 years and older (n=230,000) in France with BNT162b2 variant-adapted Fall 2023 and Spring 2024 vaccines, it is predicted that there will be 32,299 Sars-CoV-2 symptomatic infections, 3,308 hospitalization, 747 deaths, and 2,729 cases of long COVID between October 2023 – September 2024. Given the expected lower VE of the BNT162b2 variant-adapted Fall 2023 and Spring 2024 vaccines, the mRNA-1273 variant-adapted Fall 2023 and Spring 2024 vaccine is expected to prevent 3,882 infections (12%), 357 hospitalizations (11%), 81 deaths (11%), and 326 cases of long COVID compared to vaccinating with BNT162b2. The number needed to vaccinate (NNV) with the mRNA-1273 variant-adapted Fall 2023 and Spring 2024 vaccine to prevent an additional symptomatic infection, hospitalisation and death was estimated to be 118, 1,289 and 5,706, respectively. Furthermore, 645 additional QALYs would be gained with the mRNA1273 variant-adapted Fall 2023 and Spring 2024 vaccines, with over 80% of the additional QALYs gained being attributed to the prevention of death. From the health care perspective, vaccinating all IC individuals with the mRNA-1273 variant-adapted Fall 2023 and Spring 2024 vaccines would result in a €3.2 million saved in COVID-19 infection treatment costs; inclusion of productivity losses would increase cost savings to €10.1 million. Base case results are presented in Table 6.

**Table 6.** Base case results.

|  | **mRNA-1273**<br>**variant-adapted**<br>**Fall 2023 and**<br>**Spring 2024**<br>**vaccines** | **BNT162b2 variant-**<br>**adapted Fall 2023**<br>**and Spring 2024**<br>**vaccines** | **Difference\***<br>**(NNV)** |
| --- | --- | --- | --- |
| **Clinical Outcomes** |  |  |  |
| Number of vaccinations (Fall 2023 and Spring 2024) | 460,000 | 460,000 | 0 |
| **Cases** |  |  |  |
| Symptomatic infections | 28,417 | 32,299 | -3,882<br>(118) |
| Hospitalizations | 2,951 | 3,308 | -357<br>(1,289) |
| COVID-19 related deaths | 667 | 747 | -81<br>(5,706) |
| Long COVID | 2,403 | 2,729 | -326<br>(1,410) |
| **QALYs lost** |  |  |  |
| Morbidity | 417 | 472 | -54 |
| Mortality | 4,886 | 5,476 | -591 |
| **Total QALYs lost** | **5,303** | **5,948** | **-645** |
| **Economic Outcomes** |  |  |  |
| Short-term infection† | 23,253,294 € | 26,097,152 € | -2,843,858 € |
| Long COVID | 2,334,817 € | 2,651,807 € | -316,990 € |
| **Total health care perspective** | **25,588,111 €** | **28,748,959 €** | **-3,160,848 €** |
| Productivity loss | 64,099,435 € | 71,028,435 € | -6,929,000 € |
| **Total societal perspective** | 89,687,545 € | 99,777,394 € | -10,089,848 € |

\*mRNA-1273 variant-adapted Fall 2023/Spring 2024 vaccines – BNT162b2 variant-adapted Fall 2023 and Spring 2024 vaccines
†Includes cost of infection-related myocarditis
QALY: quality-adjusted life-year; NNV: number needed to vaccinate

The scenario analyses examining lower and higher COVID-19 symptomatic infection incidences, and pessimistic, “flu-like”, and alternate Fall 2023/Spring 2024 vaccine uptake continue to predict health care cost savings and QALYs gained with the use of the mRNA-1273 variant-adapted Fall 2023 and Spring 2024 vaccines compared to the BNT162b2 variant-adapted Fall 2023 and Spring 2024 vaccines. Compared to base case values, using a lower incidence of COVID-19 symptomatic infections due to a decrease in natural immunity waning resulted in an approximately 40% decrease in both cost savings and QALYs gained by the mRNA-1273 variant-adapted vaccines compared to the BNT162b2 variant-adapted vaccines. Increasing/decreasing incidence by 25%, resulted in a proportional increase/decrease in cost savings and QALYs by 25%, respectively. The alternate coverage scenario where only 50% of the IC population received the Spring 2024 vaccine had low impact on cost savings and QALYs gained (<1%). The analyses for the “flu-like” coverage scenarios resulted in cost savings and QALY gains decreases of 55%, while the pessimistic scenario resulted in cost savings and QALY gains decreases of 83%.

Sensitivity analyses results for cost savings and QALYs gained with the mRNA-1273 variant-adapted Fall 2023/Spring 2024 vaccines compared to the BNT162b2 variant-adapted Fall 2023/Spring 2024 vaccines are presented as tornado diagrams in Figure 4. Cost savings and QALYs gained were most sensitive to parameters surrounding the initial VE and waning. Varying the RR between the mRNA-1273 and BNT162b2 variant-adapted vaccines varied the initial VE of the BNT162b2 variant-adapted vaccine while keeping the initial VE of the mRNA-1273 variant-adapted vaccine the same. Using the RR from the Kopel et al. analysis decreased the difference between the VEs against hospitalization of the two vaccines. Varying costs and QALY inputs had little impact on overall results.

**Figure 3.**
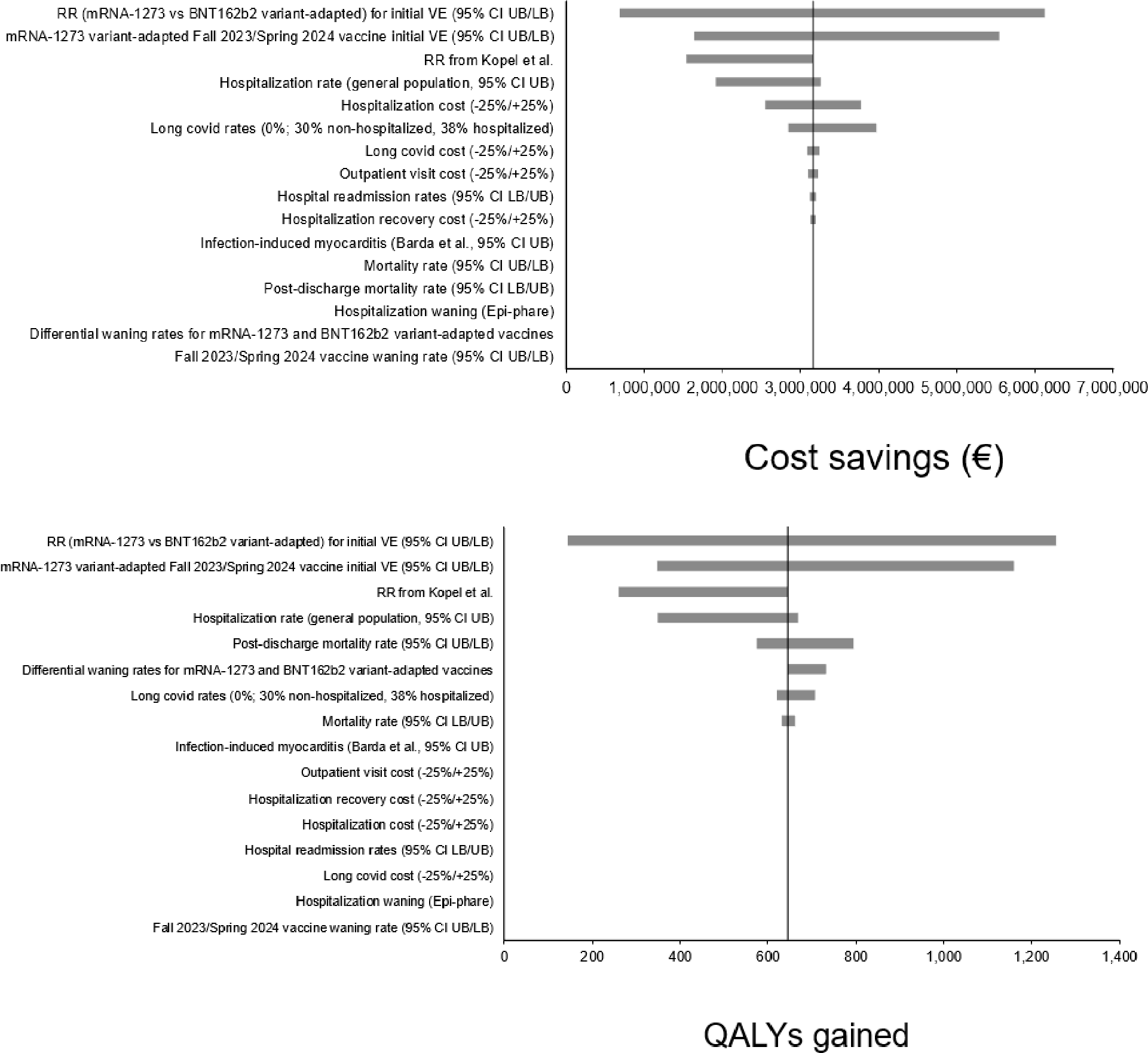
Sensitivity analyses tornado diagrams: costs and QALYs. CI: confidence interval; UB: upper bound; LB: lower bound; RR: relative risk; ICU: intensive care unit; QALY: quality-adjusted life-year

## DISCUSSION

This study examines the potential benefits associated with vaccination of the IC population aged 30 years and over in France with mRNA-1273 instead of BNT162b2 variant-adapted vaccines. As vaccination recommendations in this high-risk population are similar in Spring 2024 as in Spring 2023 [47, 69], it was assumed that the IC population received a vaccine in Fall 2023 (October 2023), and again in Spring 2024 (April 2024).

Based on prior versions of the mRNA-1273 vaccine compared to prior versions of the BNT162b2 vaccine, and using the assumption that vaccines are well-matched to the circulating variant during the 12-month period beginning October 2023, the mRNA-1273 variant-adapted Fall 2023 and Spring 2024 vaccines are assumed to have higher VEs compared to the BNT162b2 variant-adapted Fall 2023 and Spring 2024 vaccines. Vaccinating the entire IC population aged 30 years and over in October 2023 and again in April 2024 with the mRNA-1273 variant-adapted vaccines compared to the BNT162b2 variant-adapted vaccines is predicted to prevent an additional 3,882 symptomatic infections, 357 hospitalizations, 81 deaths, and 326 cases of long COVID resulting in €3.2 million saved in health care treatment costs, and 645 QALYs gained. If productivity loss is included, cost savings are predicted to reach €10.1 million (3 times the initial estimate). To contextualize this amount, these annual cost-savings would be equivalent to the treatment cost of more than 1,500 patients with cancer in France, considering a mean annual reimbursed cost of €6,289 per cancer patient [70]. Thus, these cost-savings could partly be re-allocated to French hospital systems to improve IC patients care by treating other causes of IC hospitalisation, or be used to treat other patients requiring hospital care. This is noteworthy in a context where during the COVID-19 pandemic, there was considerable strain on hospitalization capacity, which persists now amid a triple epidemic (influenza, bronchiolitis, and COVID-19).

To our knowledge, this is the first study assessing the clinical and economic impact of two mRNA variant-updated vaccines preferentially recommended for COVID-19 vaccination in France. This is also the first French health economic analysis on COVID-19 vaccines conducted in a specific subgroup of high-risk patients such as the IC patients. This study contributes to characterize COVID-19 burden in France and highlight the immediate and long-term costs associated with COVID-19 vaccination in the most vulnerable patients. Another strength of this analysis is the use of a model that allowed for the simulation of different vaccine coverage scenarios, notably akin to those observed in epidemic diseases. Study findings show that high vaccination coverage rates drive the highest public health benefits and cost-savings, which emphasizes the need for stringent vaccination campaigns and urge public health policies and healthcare professionals (including specialists) to actively promote COVID-19 vaccination during each campaign, considering the current low and insufficient coverage rates. Moreover, such analyses can help to inform and shape health policy programs for upcoming seasons, since they highlight the significant annual costs that could be saved over the years, and re-allocated to support optimization of IC patient care.

The model used for this analysis is a simplification of the real world. Although a dynamic SEIR model has been developed to compare the impact of vaccination for COVID-19 [34] and accounts for indirect protection in the population due to decreasing transmission by preventing infections, the IC population is a very small sub-set of the entire population, and the impact of vaccinating these individuals is unlikely to impact transmission in the general population [71, 72]. Therefore, this simplified static model is suitable for the population analyzed.

There is high uncertainty around future variants and the protective level of the Fall 2023 and Spring 2024 vaccines as the SARS-CoV-2 virus evolves. However, estimations of VE were based on real-world data from previous mRNA-1273 and BNT162b2 versions of the vaccines. Fall 2023 vaccines have also demonstrated robust immunologic responses that were in the same magnitude as previous formulations that were well-matched to the intended variant and subsequent circulating variants [73, 74]. Additionally, although both mRNA-1273 and BNT162b2 COVID-19 vaccines and their subsequent variant-adapted vaccines are based on mRNA technology, the formulations of mRNA-1273 versions, including their dosages and nanoparticle mRNA delivery systems, differ. As real-world studies have consistently found mRNA-1273 formulations to have higher VEs than BNT162b2 formulations [15–20], it is expected that this higher protection will remain with future versions of both vaccines, resulting in fewer infections, hospitalizations and deaths, costs savings and QALYs gained with the mRNA-1273 formulation. Sensitivity analyses found variations around VEs and waning (which affects the VE) to be the main drivers of cost savings and QALYs gained. Yet, in each analysis, vaccinating with mRNA-1273 instead of BNT162b2 variant-adapted vaccines was still generating cost savings and QALYs gained.

There is also high uncertainty surrounding the future incidence of COVID-19 infections, especially in how future waves may become seasonal. However, to date, COVID-19 prevention still requires at least one vaccination campaign every year. Therefore, advocating for health policies involves investigating all possible scenarios, as we did through multiple analyses. Furthermore, the incidence of COVID-19 was predicted using a SEIR model, and takes into account historical incidence as well as level of protection of the population, based on natural immunity and prior vaccinations. Current adapted vaccines have shown effectiveness against new and most circulating variants, including the JN.1 variant [40], which is currently the most predominant variant (>80%) circulating in France (January 2024) [75]. All incidence sensitivity analyses were consistent with the base case results, showing cost savings and QALYs gained with the use of mRNA-1273 instead of BNT162b2 variant-adapted vaccines.

IC-specific data for some of the model inputs were lacking. However, approaches were taken to minimize this impact by adjusting general population hospitalization and mortality rates using data that compared IC and non-IC cohorts. Still, not all data were adjusted. As VE data against infection were not available for mRNA-1273 formulations in the IC population, the VE for the mRNA-1273 variant-adapted Fall 2023 and Spring 2024 vaccine was assumed to be the same as the monovalent mRNA-1273 against BA.1/BA.2 [39]. As this value was based on the general population, it is likely an overestimate of the VE in IC individuals. However, as the current analysis is comparative, using a higher VE for the mRNA-1273 variant-adapted vaccine is also a conservative approach. The VE for the BNT162b2 variant-adapted vaccine is calculated by applying the RR of infection from Wang et al., (2023) [20] to the VE for variant-adapted mRNA-1273 vaccine. Mathematically, applying the RR of infection to a lower VE would result in a larger absolute VE difference between the two vaccines compared to applying the same RR to a higher VE.

Although there is emerging evidence on the protective effect of COVID-19 vaccination against long COVID, existing studies do not differentiate between the protective effect against infection and long COVID. It is unclear if the benefits seen from vaccination are only due to the avoidance of infection [76–78]. For this reason, the model did not include VE against long COVID. Instead, rates of long COVID in patients hospitalised/not hospitalised during acute infection were considered to calculate the number of long COVID episodes. Long COVID rate in those that did not require hospitalization (8%) during acute infection was obtained from Santé Publique France [54], and it was conservatively assumed the double of this rate (16%) in hospitalized patients, although rates have been previously reported as high as 38% [68]. This approach is conservative since the model does not include potential protective effect of vaccines against long COVID that would be unrelated to prevention of COVID-19 infections and COVID-19 hospitalizations, which could underestimate the clinical and economic impact of COVID-19 vaccination.

The monthly waning rates used in the analyses were based on a meta-analysis of values in the general population. In both the period of Delta and Omicron predominance, protection against hospitalization has been observed to be higher in the IC population compared to the general population [79]. Still, varying the waning rate had negligible impact on cost savings or QALYs gained, as both Moderna and Pfizer-BioNTech vaccines are waned at the same rate. Increasing the waning rate will, however, have a greater effect on the impact of only administering the Fall 2023 vaccines versus both Fall 2023 and Spring 2024 vaccines.

The base case analysis assumes 100% vaccine coverage rates. The “flu-like” and “pessimistic” coverage scenarios illustrated that cost-savings and QALYs gained decreases significantly with decreased vaccine coverage. These results highlight the importance to increase and maintain COVID-19 vaccine coverage rates high, especially in high-risk individuals, in order to prevent cases of infections and downstream consequences. Additionally, current HAS guidelines recommend not only that the IC individual receive COVID-19 vaccinations, but that their household members receive vaccination as well. This study did not examine the clinical or economic impact of household members, the impact of inclusion of these additional members on model results is unknown [35].

The QALY decrements applied to the model were based on the general population, due to lack of data in IC individuals. However, sensitivity analyses showed that varying these inputs had minimal impact on analyses results. Likewise, the proportion of hospitalized individuals requiring ICU treatment was based on the general population. It is possible that a higher rate of IC individuals requires ICU [80], resulting in higher treatment costs. Still, the model inputs are conservative, as higher ICU rates would result in greater cost savings and QALYs gained.

Similarly, resource use and costs inputs may be underestimated, as they were obtained from the general population. Preliminary data in the US IC and non-IC populations found that IC COVID-19 patients’ inpatient costs to be 1.4-fold higher than their non-IC counterparts [81]. Again, using general population data is a conservative approach as the BNT162b2 variant-adapted Fall 2023 and Spring 2024 vaccines are expected to result in more hospitalizations than the mRNA-1273 variant-adapted Fall 2023 and Spring 2024 vaccines.

In conclusion, this study illustrates the benefits of optimizing COVID-19 vaccination programmes in the IC population. According to our analyses, vaccinating the entire IC population aged 30 years and above with the mRNA-1273 variant-adapted Fall 2023 and Spring 2024 vaccine is associated with a significant public health impact and would generate €10.1 million in cost-savings (including healthcare costs and productivity losses), as well as over 645 QALYs gained over a one-year time horizon. Indeed, maximization of vaccination coverage rates and preferential use of vaccines offering the highest level of protection to the most vulnerable patients will have a significant impact on reducing infections and downstream clinical and economic outcomes. Yet, having both vaccines available in France will allow for the option to protect more infection and hospitalization in a vulnerable population, potentially improving patient flow in hospital settings during the autumn and winter seasons, which are conducive to respiratory tract infections.

## Author contributions

Conceptualization, AL, MK, MM, EB, KJ, NE, BD; Methodology, AL, MM, MK; Software, MM; Validation, AL and MK; Formal analysis, AL; Writing – original draft preparation, AL and MK; Writing – review & editing, AL, MM, MK, BD, CD, EB, KJ, NE, MU; Funding Acquisition: EB

## Data availability statement

The datasets used and/or analyzed during the current study are available from the corresponding author on reasonable request.

## Supporting information

Technical Appendix

## Acknowledgements

Not applicable

