## Supplementary material for "Substantial reduction in the clinical and economic burden of disease following variant-adapted mRNA COVID-19 vaccines in immunocompromised patients in France": Technical Appendix

**TITLE:**

### Adaptation of the US SEIR model

The following parameters have been adapted from the US Susceptible-Exposed-Infected-Recovered (SEIR) model[1] to France using French-specific inputs:

- Population size[2]
- Contact matrix/mixing pattern[3]
- Social distancing and mask use[4]
- Historical vaccine coverage[5]
- Vaccine effectiveness based on market shares of prior vaccines administered[5]

The total number of infections in the population predicted by the SEIR model was calibrated to French-specific targets (daily incidence of reported COVID-19 cases between February 4, 2020-May 31, 2022) from the Institute for Health Metrics and Evaluation (IHME)[6].

The adapted SEIR model was used to generate residual vaccine effectiveness (VE) inputs and symptomatic incidence rates for the static model used for the immunocompromised (IC) population.

### Residual vaccine effectiveness

Although not used in the base case analysis, as 100% of the cohort receives a Fall 2023 vaccine in Month 1 of the analytic time horizon, residual VE is necessary to estimate the level of protection in the population entering the model at the start of the analytic time horizon (October 1, 2023) due to prior vaccinations if the entire cohort does not receive a vaccine in Month 1. The France-adapted SEIR model is used to calculate the vaccine-induced protection remaining the in population, taking into consideration the timing of the prior boosters, VE of primary series and boosters against the dominant variant at time of administration, and the historical coverage in the IC population. These values are displayed below in Table 1 to Table 6.

Table 1. Residual vaccine effectiveness against infection – Primary series, first booster or second booster

| **Age Group** | **Month** | | | | | | | | | | | |
| --- | --- | --- | --- | --- | --- | --- | --- | --- | --- | --- | --- | --- |
|  | **1** | **2** | **3** | **4** | **5** | **6** | **7** | **8** | **9** | **10** | **11** | **12** |
| 30-39 years | 0.000 | 0.000 | 0.000 | 0.000 | 0.000 | 0.000 | 0.000 | 0.000 | 0.000 | 0.000 | 0.000 | 0.000 |
| 40-49 years | 0.000 | 0.000 | 0.000 | 0.000 | 0.000 | 0.000 | 0.000 | 0.000 | 0.000 | 0.000 | 0.000 | 0.000 |
| 50-59 years | 0.000 | 0.000 | 0.000 | 0.000 | 0.000 | 0.000 | 0.000 | 0.000 | 0.000 | 0.000 | 0.000 | 0.000 |
| 60-64 years | 0.000 | 0.000 | 0.000 | 0.000 | 0.000 | 0.000 | 0.000 | 0.000 | 0.000 | 0.000 | 0.000 | 0.000 |
| 65-69 years | 0.000 | 0.000 | 0.000 | 0.000 | 0.000 | 0.000 | 0.000 | 0.000 | 0.000 | 0.000 | 0.000 | 0.000 |
| 70-79 years | 0.000 | 0.000 | 0.000 | 0.000 | 0.000 | 0.000 | 0.000 | 0.000 | 0.000 | 0.000 | 0.000 | 0.000 |
| 80+ years | 0.000 | 0.000 | 0.000 | 0.000 | 0.000 | 0.000 | 0.000 | 0.000 | 0.000 | 0.000 | 0.000 | 0.000 |

Table 2. Residual vaccine effectiveness against infection – third booster

| **Age Group** | **Month** | | | | | | | | | | | |
| --- | --- | --- | --- | --- | --- | --- | --- | --- | --- | --- | --- | --- |
|  | **1** | **2** | **3** | **4** | **5** | **6** | **7** | **8** | **9** | **10** | **11** | **12** |
| 30-39 years | 0.034 | 0.000 | 0.000 | 0.000 | 0.000 | 0.000 | 0.000 | 0.000 | 0.000 | 0.000 | 0.000 | 0.000 |
| 40-49 years | 0.034 | 0.000 | 0.000 | 0.000 | 0.000 | 0.000 | 0.000 | 0.000 | 0.000 | 0.000 | 0.000 | 0.000 |
| 50-59 years | 0.036 | 0.000 | 0.000 | 0.000 | 0.000 | 0.000 | 0.000 | 0.000 | 0.000 | 0.000 | 0.000 | 0.000 |
| 60-64 years | 0.043 | 0.000 | 0.000 | 0.000 | 0.000 | 0.000 | 0.000 | 0.000 | 0.000 | 0.000 | 0.000 | 0.000 |
| 65-69 years | 0.043 | 0.000 | 0.000 | 0.000 | 0.000 | 0.000 | 0.000 | 0.000 | 0.000 | 0.000 | 0.000 | 0.000 |
| 70-79 years | 0.040 | 0.000 | 0.000 | 0.000 | 0.000 | 0.000 | 0.000 | 0.000 | 0.000 | 0.000 | 0.000 | 0.000 |
| 80+ years | 0.029 | 0.000 | 0.000 | 0.000 | 0.000 | 0.000 | 0.000 | 0.000 | 0.000 | 0.000 | 0.000 | 0.000 |

Table 3. Residual vaccine effectiveness against hospitalization – Primary series

| **Age Group** | **Month** | | | | | | | | | | | |
| --- | --- | --- | --- | --- | --- | --- | --- | --- | --- | --- | --- | --- |
|  | **1** | **2** | **3** | **4** | **5** | **6** | **7** | **8** | **9** | **10** | **11** | **12** |
| 30-39 years | 0.167 | 0.149 | 0.133 | 0.115 | 0.098 | 0.081 | 0.064 | 0.047 | 0.030 | 0.013 | 0.000 | 0.000 |
| 40-49 years | 0.167 | 0.149 | 0.133 | 0.115 | 0.098 | 0.081 | 0.064 | 0.047 | 0.030 | 0.013 | 0.000 | 0.000 |
| 50-59 years | 0.161 | 0.144 | 0.127 | 0.109 | 0.092 | 0.076 | 0.058 | 0.041 | 0.024 | 0.007 | 0.000 | 0.000 |
| 60-64 years | 0.156 | 0.139 | 0.122 | 0.104 | 0.087 | 0.071 | 0.053 | 0.036 | 0.019 | 0.002 | 0.000 | 0.000 |
| 65-69 years | 0.156 | 0.139 | 0.122 | 0.104 | 0.087 | 0.071 | 0.053 | 0.036 | 0.019 | 0.002 | 0.000 | 0.000 |
| 70-79 years | 0.146 | 0.129 | 0.112 | 0.095 | 0.077 | 0.061 | 0.043 | 0.027 | 0.009 | 0.000 | 0.000 | 0.000 |
| 80+ years | 0.141 | 0.123 | 0.107 | 0.089 | 0.072 | 0.055 | 0.038 | 0.021 | 0.004 | 0.000 | 0.000 | 0.000 |

Table 4. Residual vaccine effectiveness against hospitalization – First booster

| **Age Group** | **Month** | | | | | | | | | | | |
| --- | --- | --- | --- | --- | --- | --- | --- | --- | --- | --- | --- | --- |
|  | **1** | **2** | **3** | **4** | **5** | **6** | **7** | **8** | **9** | **10** | **11** | **12** |
| 30-39 years | 0.299 | 0.284 | 0.270 | 0.256 | 0.242 | 0.228 | 0.214 | 0.200 | 0.186 | 0.172 | 0.158 | 0.143 |
| 40-49 years | 0.299 | 0.284 | 0.270 | 0.256 | 0.242 | 0.228 | 0.214 | 0.200 | 0.186 | 0.172 | 0.158 | 0.143 |
| 50-59 years | 0.296 | 0.282 | 0.268 | 0.253 | 0.239 | 0.226 | 0.211 | 0.197 | 0.183 | 0.169 | 0.155 | 0.140 |
| 60-64 years | 0.293 | 0.279 | 0.265 | 0.251 | 0.236 | 0.223 | 0.208 | 0.195 | 0.180 | 0.166 | 0.152 | 0.138 |
| 65-69 years | 0.293 | 0.279 | 0.265 | 0.251 | 0.236 | 0.223 | 0.208 | 0.195 | 0.180 | 0.166 | 0.152 | 0.138 |
| 70-79 years | 0.288 | 0.274 | 0.260 | 0.246 | 0.231 | 0.218 | 0.204 | 0.190 | 0.175 | 0.162 | 0.147 | 0.133 |
| 80+ years | 0.285 | 0.271 | 0.257 | 0.243 | 0.228 | 0.215 | 0.201 | 0.187 | 0.172 | 0.158 | 0.144 | 0.130 |

Table 5. Residual vaccine effectiveness against hospitalization – Second booster

| **Age Group** | **Month** | | | | | | | | | | | |
| --- | --- | --- | --- | --- | --- | --- | --- | --- | --- | --- | --- | --- |
|  | **1** | **2** | **3** | **4** | **5** | **6** | **7** | **8** | **9** | **10** | **11** | **12** |
| 30-39 years | 0.658 | 0.644 | 0.630 | 0.616 | 0.601 | 0.588 | 0.573 | 0.560 | 0.545 | 0.531 | 0.517 | 0.503 |
| 40-49 years | 0.658 | 0.644 | 0.630 | 0.616 | 0.601 | 0.588 | 0.573 | 0.560 | 0.545 | 0.531 | 0.517 | 0.503 |
| 50-59 years | 0.653 | 0.638 | 0.625 | 0.610 | 0.596 | 0.582 | 0.568 | 0.554 | 0.540 | 0.526 | 0.512 | 0.497 |
| 60-64 years | 0.658 | 0.644 | 0.630 | 0.616 | 0.601 | 0.588 | 0.574 | 0.560 | 0.545 | 0.532 | 0.517 | 0.503 |
| 65-69 years | 0.658 | 0.644 | 0.630 | 0.616 | 0.601 | 0.588 | 0.574 | 0.560 | 0.545 | 0.532 | 0.517 | 0.503 |
| 70-79 years | 0.625 | 0.611 | 0.597 | 0.582 | 0.568 | 0.555 | 0.540 | 0.526 | 0.512 | 0.498 | 0.484 | 0.469 |
| 80+ years | 0.528 | 0.513 | 0.499 | 0.485 | 0.471 | 0.457 | 0.443 | 0.429 | 0.415 | 0.401 | 0.387 | 0.372 |

Table 6. Residual vaccine effectiveness against hospitalization – Third booster

| **Age Group** | **Month** | | | | | | | | | | | |
| --- | --- | --- | --- | --- | --- | --- | --- | --- | --- | --- | --- | --- |
|  | **1** | **2** | **3** | **4** | **5** | **6** | **7** | **8** | **9** | **10** | **11** | **12** |
| 30-39 years | 0.741 | 0.726 | 0.712 | 0.698 | 0.684 | 0.670 | 0.656 | 0.642 | 0.628 | 0.614 | 0.600 | 0.585 |
| 40-49 years | 0.741 | 0.726 | 0.712 | 0.698 | 0.684 | 0.670 | 0.656 | 0.642 | 0.628 | 0.614 | 0.600 | 0.585 |
| 50-59 years | 0.741 | 0.727 | 0.713 | 0.699 | 0.684 | 0.671 | 0.657 | 0.643 | 0.628 | 0.615 | 0.600 | 0.586 |
| 60-64 years | 0.743 | 0.729 | 0.715 | 0.701 | 0.686 | 0.673 | 0.659 | 0.645 | 0.630 | 0.616 | 0.602 | 0.588 |
| 65-69 years | 0.743 | 0.729 | 0.715 | 0.701 | 0.686 | 0.673 | 0.659 | 0.645 | 0.630 | 0.616 | 0.602 | 0.588 |
| 70-79 years | 0.742 | 0.728 | 0.714 | 0.700 | 0.685 | 0.672 | 0.658 | 0.644 | 0.629 | 0.616 | 0.601 | 0.587 |
| 80+ years | 0.739 | 0.725 | 0.711 | 0.697 | 0.682 | 0.669 | 0.655 | 0.641 | 0.626 | 0.613 | 0.598 | 0.584 |

### Scenario analysis: Incidence of symptomatic infection

The SEIR model was also used to project a lower cumulative incidence of symptomatic COVID-19 infection for a scenario analysis. In brief, in the SEIR model, the waning of naturally acquired immunity was assumed to be slower (half of base case rate = 1.85%) than in the base case (assumed to be the same as waning of infection against primary series vaccination for BA.1/BA.2), or in other words, the mean duration of natural immunity was assumed to be longer. In the dynamic SEIR model, this assumption leads to people remaining in the R (Recovered) state for a longer period of time. This in turn reduces the overall number of people in the S (Susceptible) state and reduces the force of infection, leading to a lower projected annual incidence in the French population. Inputs are displayed in Table 7. Additional scenario analyses were also conducted where the incidence was increased/decreased by 25% of the base case values.

Table 7. Scenario analysis: projected monthly incidence of symptomatic COVID-19 infection amongst the unvaccinated in France (%)

| **Age Group** | **Month*** | | | | | | | | | | | |
| --- | --- | --- | --- | --- | --- | --- | --- | --- | --- | --- | --- | --- |
|  | **1** | **2** | **3** | **4** | **5** | **6** | **7** | **8** | **9** | **10** | **11** | **12** |
| 30-39 years | 2.47 | 2.58 | 1.80 | 1.04 | 0.64 | 0.53 | 0.48 | 0.54 | 0.62 | 0.83 | 1.09 | 1.35 |
| 40-49 years | 2.45 | 2.57 | 1.81 | 1.05 | 0.64 | 0.53 | 0.48 | 0.53 | 0.62 | 0.82 | 1.08 | 1.33 |
| 50-59 years | 2.24 | 2.57 | 1.94 | 1.13 | 0.67 | 0.53 | 0.46 | 0.49 | 0.55 | 0.73 | 0.96 | 1.21 |
| 60-64 years | 2.01 | 2.58 | 2.19 | 1.33 | 0.76 | 0.56 | 0.44 | 0.45 | 0.48 | 0.61 | 0.79 | 1.01 |
| 65-69 years | 2.01 | 2.58 | 2.19 | 1.33 | 0.76 | 0.56 | 0.44 | 0.45 | 0.48 | 0.61 | 0.79 | 1.01 |
| 70-79 years | 1.92 | 2.53 | 2.21 | 1.37 | 0.77 | 0.56 | 0.44 | 0.43 | 0.46 | 0.58 | 0.75 | 0.95 |
| ≥80 years | 1.84 | 2.43 | 2.12 | 1.32 | 0.75 | 0.55 | 0.43 | 0.42 | 0.45 | 0.56 | 0.73 | 0.93 |

*Month 1 = October 2023.

### Productivity loss costs

Costs from the societal perspective include all direct health care costs due to COVID-19 infection treatment, as well as costs due to productivity loss due to vaccine administration, acute infection and hospitalization, and long COVID. The model includes productivity loss costs due to acute infection and hospitalizations.

The number of days lost per infection, hospitalization and long COVID are provided in Table 8. They were multiplied by the average daily wage, and adjusted for the labour force participation rate, by age.

Table 8. Lost productivity

| **Model Parameter** | **Value** | **Data Source** |
| --- | --- | --- |
| Labour force participation rate |  |  |
| 0-17 years | 8% | INSEE (2022)[7] |
| 18-29 years | 61% |  |
| 30-39 years | 88.3% |  |
| 40-49 years | 88.3% |  |
| 50-59 years | 79.3% |  |
| 60-64 years | 69.9% |  |
| 65-69 years | 0.0% |  |
| 70-79 years | 0.0% |  |
| 80+ years | 0.0% |  |
| Average Wage: GDP per person per working day | 351.13€ | INSEE (2022)[7, 8]; Capeos (2022)[9] |
| Work Days Lost for: | | |
| Vaccine administration | 0.18 days | Prosser et al. (2019)[10] |
| Infection-induced myocarditis | 3 days | HCUP[11] |
| Infection: not hospitalized | 7 | Recommendation of the Ministry of Health to isolate for 7 days[12] |
| Infection: hospitalized |  |  |
| General ward | 10.4 days | ScanCovid, 2022[13] |
| ICU | 18.9 days | ScanCovid, 2022[13] |
| Hospitalization recovery | 33.43 days | Chopra et al. (2021)[14] |
| Long COVID | 32.47 days | Davis et al. (2021)[15] |

INSEE: Institute National e la Statistique et des Études Économique; HCUP: Healthcare Cost and Utilization Project; GDP: Gross domestic product; ICU: Intensive care unit

### List of sensitivity analyses

Deterministic sensitivity analyses and values varied for each analysis are displayed in

Table 9. Where available, 95% confidence intervals were used. All other values were varied by ±25%.

Table 9. List of deterministic sensitivity analyses

| **Parameter** | **Base Case** | **Range** |
| --- | --- | --- |
| mRNA-1273 variant-adapted Fall 2023 and Spring 2024 vaccine initial VE | Tseng et al., for hospitalization,[16] Pratama et al., for infection[17] | 95% CI |
| RR between mRNA-1273 and BNT162b2 variant-adapted Fall 2023 and Spring 2024 vaccines | Wang et al.,[18] | 95% CI |
| Relative VE between mRNA-1273 and BNT162b2 variant-adapted Fall 2023 and Spring 2024 vaccines | Wang et al.,[18] | 15.0% (infection); 4.8% (hospitalization) |
| Waning (Fall 2023 and Spring 2024 vaccine) | Higdon et al.,[19] | 95% CI |
| Differential waning between mRNA-1273 and BNT162b2 variant-adapted Fall 2023 and Spring 2024 vaccines | Higdon et al.,[19] | RR for infection and hospitalization from Wang et al.,[18] maintained between the mRNA-1273 and BNT162b2 variant-adapted vaccines for the entire analytic time horizon by adjusting the waning rates for the BNT162b2 variant-adapted vaccine |
| Hospitalization waning rate | Higdon et al.,[19] | Epi-phare data[20] |
| Hospitalization rate | RR for IC population applied to general population hospitalization rates | 95% CI[21] |
|  |  | General population hospitalization rates[22] |
| Mortality rate |  | 95% CI |
| Hospital readmission rate | 4.1%[23] | 95% CI[23] |
| Post discharge mortality rate | 2.7%[23] | 95% CI[23] |
| Long COVID rates | 8%[24] | 0% (no long COVID) |
|  |  | 30% non-hospitalized; 38% hospitalized[25] |
| Infection-induced myocarditis | Boehmer et al.,[26] | Barda et al.,[27] |
| Cost of ambulatory care per patient, estimated based on:  -Unit cost of home care withou**t** oxygen therapy and symptomatic treatment (including 2 GP visits, 1 RT-PCR test, 2 antigenic tests, paracetamol and NSAID): 148.67 €  -Unit cost of home care with oxygen therapy and symptomatic treatment (including 2 GP visits, 1 cardiologist visit, 7 nurse home visits, 1 physiotherapist visit, paracetamol, NSAID, oxygen therapy, 1 radiology exam, 1 RT-PCR test, 2 antigenic tests):1,009.06 €  -Percentage of patients with Covid 19 severe symptoms requiring oxygen therapy at home: 15%  (0,85 % * 148,67 + 0,15 % * 1,009.06 = 277.73). | 277.73 €[28-30] | ±25% |
| Cost per hospitalization, estimated based on:  -Unit cost of ‘No ICU or ventilator’ hospital stay, estimated according to the unit cost and percentage of patients hospitalised at home or in general wards:  5.56% * 1,963.04 + 94.44% * 4935.23 = 4770.11€  ‘ICU stay’ unit cost: 17,093€ | No ICU or ventilator: 4,770€[31, 32]  ICU: 17,093€[31, 32] | ±25% |
| Cost per hospitalization in follow-up and rehabilitation unit, estimated based on:  -Unit cost of follow-up and rehabilitation unit (including transport cost)  -Percentage of patients requiring follow-up and rehabilitation: 10% | 474€[31-33] | ±25% |
| Cost of long COVID (over a time period of 6 months: including 2 GP visits, 1 physiotherapist visit per week, 3 specialist visits, 2 PET scan, 1 echocardiography, paracetamol, NSAID, antidepressant) | 971.71€ [28, 29, 34] | ±25% |
| Short term infection QALY decrement | 0.0036[35] | ±25% |
| Hospitalization QALYs decrement | No ICU or ventilator stay: 0.022;[36]  ICU stay: 0.466[36] | ±25% |
| Long COVID QALYs decrement | Non-hospitalized: 0.069;[37]  Hospitalized: 0.139[37] | ±25% |

CI, confidence interval, VE, vaccine effectiveness; RR, relative risk; ICU, intensive care unit; QALY, quality-adjusted life-year; GP, general practitioner; RT-PCR, reverse transcriptase polymerase chain reaction; NSAID, non-steroidal anti-inflammatory drug; PET, positron emission tomography.

### Sensitivity analyses results

Additional results from the sensitivity analyses (symptomatic infections, hospitalizations, and deaths prevented) are displayed as tornado diagrams below in Figure 1.

Figure 1. Sensitivity analyses: Tornado diagrams for infections, hospitalizations, and deaths prevented with mRNA-1273 variant-adapted Fall 2023 and Spring 2024 vaccines compared to BNT162b2 variant-adapted Fall 2023 and Spring 2024 variant-adapted vaccines

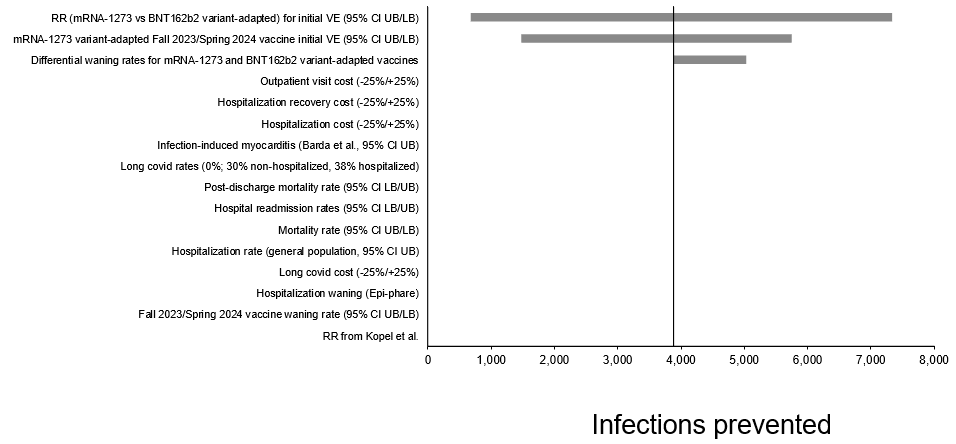

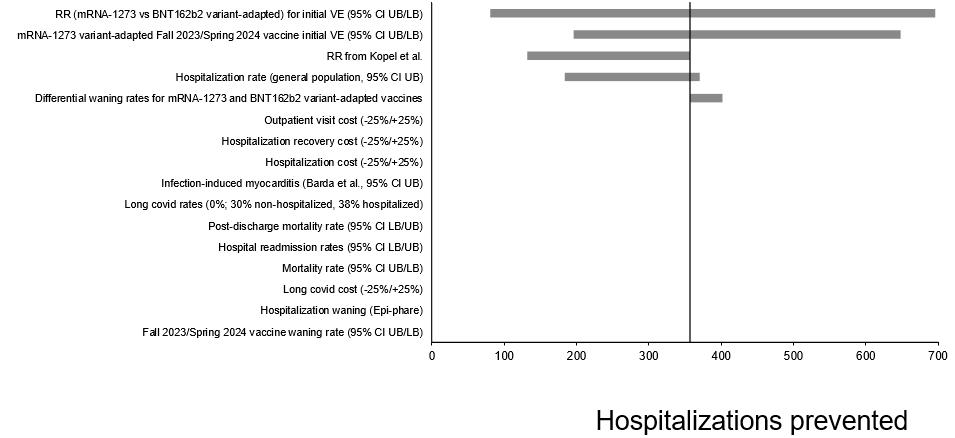

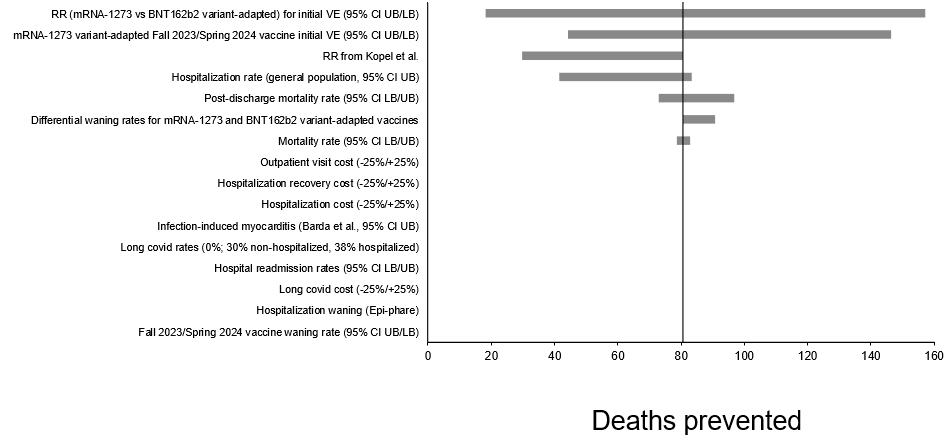

CI: confidence interval; UB: upper bound; LB: lower bound; RR: relative risk; ICU: intensive care unit; QALY: quality-adjusted life-year

12.	Gouvernement de France. *Informations Coronavirus.*; Available from: <https://www.gouvernement.fr/info-coronavirus>.

30.	EPI-PHARE. *Recours à l’oxygénothérapie à domicile pour une infection à SARS-CoV-2 en 2021.* . 2022.

31.	Gallien, S., et al., *Couts des hospitalisations et des soins de suite et de réadaptation liés au COVID-19 en France en 2020. Médecine et Maladies Infectieuses Formation 2022: S51*. 2022.

32.	Cour des comptes. *Rapport sur l'application des lois de financement de la sécurité sociale*. 2019 October 8, 2019 [cited 2023 March 1]; Available from: Rapport sur l'application des lois de financement de la sécurité sociale.
